# Single-Center Randomized Controlled Trial of Middle Meningeal Artery Embolization for Chronic Subdural Hematoma

**DOI:** 10.1101/2025.04.28.25323504

**Authors:** Armin D. Tavakkoli, Julio D. Montejo, Katelyn E. Salotto, Zabiullah Bajouri, William J. Smith, Caitlin A. Payne, James E. Lee, John H. Kanter, Imad S. Khan, Vyacheslav I. Makler, Evalina Bond, Kelsey P. Kinsman, Stephanie A. Ihezie, David Soucy, Barbara C. Badgley, Amber L. Porter, Beverly A. Allen, Stephen J. Guerin, Naser Jaleel, Daniel R. Calnan, Clifford J. Eskey

## Abstract

**Background:** Embolization of the middle meningeal artery (MMA) for the treatment of chronic subdural hematoma (cSDH) has gained increasing acceptance since the publication of several nonrandomized studies that appear to demonstrate a high level of efficacy for the procedure both when performed as an adjunct to patients undergoing surgical drainage and when performed in minimally symptomatic patients treated without surgical drainage.

**Methods:** We conducted a single-center, randomized controlled trial (RCT) of MMA embolization in consecutive patients presenting to our hospital with cSDH from April 2020 to July 2023. Enrolled patients were randomized to standard of care (observation vs. surgical drainage per the treating neurosurgeon) or standard of care plus MMA embolization. The primary endpoint was the combined criteria of cSDH resolution (defined as < 5mm), resolution of clinical symptoms, no further intervention, and no major treatment-related complications.

**Results:** In total, 190 patients were screened, 46 were enrolled/randomized, and 1 withdrew immediately after randomization but before treatment. Accrual was ended early due to diminished enrollment; 79% of the accrual goal was achieved. Of the 45 study patients, 40 had sufficient follow up for primary outcome analysis. The primary endpoint was met by 14/20 (70%) in the control group and 15/20 (75%) in the MMA embolization group, respectively, p=0.5.

**Conclusions:** In this RCT, no significant difference was observed between standard of care and standard of care plus MMA embolization for cSDH. While the effect size appears to be smaller than that suggested by early case series, ongoing trials with greater statistical power may show benefit for the procedure.

**Previously presented in abstract form at:** New England Neurosurgical Society (NENS) 2024 Meeting

**Key Messages:** **What is already known on this topic** – Middle meningeal artery (MMA) embolization appears to be a safe and effective treatment for chronic subdural hematoma (cSDH), based on large case series. Randomized controlled trials of MMA embolization are just now being completed.

**What this study adds** – This study found no significant difference in the primary endpoint within 1 year (resolution of cSDH defined as < 5mm, no return of clinical symptoms, no need for further intervention, and no major treatment-related complications).

**How this study might affect research, practice or policy** – Further research, specifically RCTs with greater statistical power, are needed to better elucidate whether MMA embolization adds a statistically significant benefit to standard of care in cSDH.

## Introduction

Chronic subdural hematomas (cSDH) are one of the most common conditions encountered by neurosurgeons with an estimated incidence of 1-5.3 cases per 100,000 population ^1^. Chronic SDH is more common in elderly adults, and incidence is increasing. The neurologic impact of cSDH can range from asymptomatic to profoundly symptomatic including seizures, hemiparesis, sensory disturbances, altered consciousness, and death. Standard of care for cSDH has largely consisted of surgical drainage via either minimally invasive surgical approaches such as subdural drain placement, subdural evacuating port system (SEPS) placement, or burr hole drainage, versus open surgical approaches including mini- or full-craniotomy. These procedures are usually effective at rapidly reducing the mass effect from the hematoma.

These approaches do not treat a major component of the underlying pathology. The presence of blood in the subdural space creates an inflammatory cascade that leads to formation of inflammatory membranes with immature blood vessels. These vascular membranes produce both exudate and hemorrhage ^2^. As a result, recurrence of the hemorrhagic collection may occur. One study found a treatment failure rate of 10-32% and a need for additional surgery at 19-40%^3–5^.

To address the problems of cSDH progression and recurrence, physicians have recently employed embolization of the middle meningeal artery (MMA). Neurointerventional trained clinicians, using percutaneous, catheter-based techniques, access the middle meningeal artery and occlude the vessel with one of several embolic materials. The resulting devascularization of membranes and dura reduces further exudation and into the subdural space. A pilot study of 72 patients who underwent MMA embolization showed a 1.4% recurrence rate compared to 27.5% in the historical control group without MMA embolization ^6^. Physicians have increasingly used this procedure both as a stand-alone treatment in minimally symptomatic patients and as an adjunct treatment in patients requiring urgent subdural drainage.

Several groups of investigators are currently pursuing randomized controlled trials (RCT) of MMA embolization^1,3,5,6^. However, the published results to date are mixed and they show a smaller effect size than those obtained from case series ^7^. We performed a single-center RCT to evaluate the efficacy of MMA embolization for cSDH as an adjuvant treatment to the standard of care, whether that be surgical drainage or observation in the appropriate scenarios.

We hypothesized that the addition of MMA embolization as an adjuvant treatment to the standard of care for cSDH would improve the rate of cSDH treatment success. To test this hypothesis, we conducted a single-center randomized controlled trial of standard of care (control group) vs standard of care plus MMA embolization (intervention group) for the treatment of cSDH.

## Methods

### Design and Oversight

We conducted a single-center prospective RCT. The trial protocol was approved by the institutional IRB and registered with clinicaltrials.gov (NCT04270955) prior to any patient screening or enrollment. The manuscript was prepared in accordance with the CONSORT guidelines ^8–10^.

### Participants

#### Inclusion Criteria

Consecutive adult patients with cSDH who presented to the DHMC emergency department or inpatient wards were screened for inclusion. Included patients were 18 years of age or older, had computed tomography (CT) imaging showing a cSDH ≥ 7mm in maximal thickness encompassing ≥ 50% of the convexity (non-focal), and capability of giving consent for the procedure or have an acceptable surrogate capable of giving consent on the subject’s behalf.

#### Exclusion Criteria

Patients were excluded if imaging demonstrated an underlying vascular malformation, tumor, spontaneous cerebrospinal fluid (CSF) hypotension or previous craniotomy, life expectancy < 6 months, prior evidence of vascular anatomy that would put the patient at high risk for adverse events (e.g. critical carotid stenosis, abnormal external-internal carotid circulation), history of neurosurgical procedure ipsilateral to the cSDH, inability to follow-up, and vulnerable patients including homeless patients, incarcerated patients and mentally ill patients without appropriate medical decision-making proxy that the physician believes are incapable of appropriately assessing the risks of the procedure.

#### Enrollment

Eligible patients were offered enrollment in the trial after independent clinical assessment and standard of care recommendation (either observation or surgical drainage) by the responsible neurosurgeon. All study patients and/or appropriate surrogate decision makers agreed to proceed voluntarily and provided written informed consent prior to enrollment and randomization.

### Sample size

The sample size was determined via a power analysis based on the meta-analysis by Srivatsan et al.^14^, where cSDH treatment failure rate for symptomatic and asymptomatic patients without MMA embolization was 27.7%, with MMA embolization the failure rate was 2.1%. With an alpha error set to 0.05 and power set to 80%, the estimated sample size needed is 58 patients (with 1:1 enrollment).

### Randomization

#### Sequence generation

In advance of trial initiation, two block sheets with random allocation sequences, one for the symptomatic subgroup and another for the asymptomatic subgroup, were prepared using block randomization in groups of 4. Consecutive patients enrolled in the study were consecutively added to the block sheets thereby randomly assigning their treatment group. The block randomization kept the numbers assigned to each group random but near equal throughout the process. The random allocation sequence block sheets were not concealed to study personnel.

### Blinding

Study participants were not blinded to their group assignment. Personnel providing clinical care, including in the inpatient and outpatient settings, and procedural/surgical care were not blinded to the participant’s group assignment. The final determination, however, of whether a study participant had successful treatment as pre-defined in the protocol was made in a blinded fashion by the senior author. Masking was accomplished by presenting the senior author with objective and de-identified information that did not contain the assigned group but contained information relevant to the determination of treatment success.

### Interventions

#### Parallel Study Arms

Patients were initially categorized based on presentation: symptomatic or asymptomatic. Symptomatic patients were defined as those with a neurological symptom or deficit attributable to the cSDH including but not limited to severe headache, weakness, numbness, and alterations in consciousness. For symptomatic patients, urgent decompression had been performed or was planned. Independently within each subgroup, patients were then randomly assigned to standard of care alone (control group) vs. standard of care plus MMA embolization (intervention group).

#### Standard of Care

All patients received standard of care as determined by the responsible attending neurosurgeon. Standard of care consisted of either observation or standard hematoma evacuation by either minimally invasive surgical approaches (subdural drain placement, subdural evacuating port system placement, or burr hole) or by open surgical approaches (mini or full craniotomy) ^11–13^. The trial protocol did not specify or limit how or when standard of care was to be administered.

### MMA Embolization

#### Timing

Patients randomized to the intervention group first received MMA embolization at the earliest possible time point deemed safe by the clinical team. Every effort was made to minimize the number of days until in-trial MMA embolization was performed with a goal of no later than 2 weeks of enrollment. Preferably, the in-trial MMA embolization was performed during their index hospitalization if able, however, outpatient in-trial MMA embolization was allowed within 2 weeks from enrollment.

#### Laterality

For unilateral cSDH, MMA embolization was performed ipsilaterally. For bilateral cSDH (in patients randomized to the intervention group), all cSDHs satisfying the 7 mm threshold were embolized.

#### Procedure details

The MMA embolization procedure was performed by one of three neurointerventionalists using standard catheter-based angiography techniques^6^. Briefly, under general anesthesia either common femoral or radial artery access was obtained followed by angiography of the ipsilateral internal and external carotid arteries and middle meningeal arteries to characterize the MMA anatomy and assess for potential collateral pathways to the cerebral and central retinal arteries. If no abnormal communication was identified and embolization of the MMA was considered safe and feasible, the distal MMA branches were occluded using microparticle Embospheres (Merit Medical Systems Inc., USA). In one case with collateral supply to the orbit, a more proximal MMA embolization was performed with detachable coils (Stryker Inc, USA). The materials used and extent of embolization were determined by the performing attending physician.

#### Off-trial MMA embolization

Patients randomized to the control group who later failed their initial treatment pathway as determined by the trial protocol were eligible for off-trial MMA embolization. The timing goals were not applicable in these cases, but the laterality and procedure details were identical.

### Clinical and imaging follow-up after hospital discharge

Patients were seen in Neurosurgery clinic, either in person or via video telehealth, at approximately 3 months, 6 months, and 1 year after discharge from the hospital. These assessments were performed by clinical staff who were not part of the research team. The assessments included NIHSS, mRS, and evaluation for adverse events. CT scans were obtained at each of these time points unless the cSDH had resolved at an earlier time point.

### Outcomes

#### Primary Outcomes

The primary endpoint was treatment success as defined by a patient meeting all the composite criteria: (a) complete or near complete resolution of the cSDH (defined as <5 mm in maximal coronal thickness) within 1 year of enrollment (b) absence of recurrent clinical symptoms attributable to the hematoma, (c) absence of major adverse procedure-related events, and (d) absence of additional open surgical or endovascular intervention for the cSDH. Major adverse procedure-related events included but not limited to death, stroke, parenchymal hemorrhage, new neurological deficit, and symptomatic deterioration resulting in increased level of care.

Assessment of study CT scans was performed in a blinded-fashion by a board-certified neuro-radiologist who evaluated images without knowledge of the subject’s study group assignment or clinical data. Determination of the primary and secondary endpoint classifications were performed by consensus among authors JDM/ADT/CJE.

#### Secondary Outcomes

Secondary outcomes were (a) survival at 1 year after enrollment as determined by all-cause mortality independent of whether the primary endpoint had been met at a previous time point, (b) NIHSS relative to baseline at 3 months, 6 months, and 1 year, and (c) mRS relative to baseline at 3 months, 6 months, and 1 year.

#### Statistical methods

Groups for analysis were determined by intention-to-treat. Outcome analyses, including the comparisons of treatment success between the control and the intervention groups, were performed using Fisher’s exact test. The effect of study arm (symptomatic vs. asymptomatic) was examined using logistic regression. In order to assess the effect of embolization on the timing of the outcome variables, time-to-event analysis was performed for treatment failure and, separately, for resolution of cSDH on imaging.

All analyses were performed using R Statistical Software (R Core Team, v. 4.4.2).

## Results

### Recruitment

The trial initially began in February 2020. Due to the COVID-19 pandemic, the study was halted in March 2020. To maintain the consecutive integrity of the trial, the decision was made at that time to exclude patients screened and enrolled prior to the trial halt. Once the status of the COVID-19 pandemic allowed us to resume the study, the trial was then restarted in April 2020 which is considered the formal start date of this trial. Enrollment continued for over 3 years until July 2023.

In total, 190 consecutive patients were screened, 124 met criteria for enrollment. In a few situations encountered during the course of the study, eligible patients could not be offered enrollment due to 1) the treating neurosurgeon requested MMA embolization be performed off trial (n=9), 2) there was a gap in neuro-interventional coverage (n=2), and 3) the patient left the hospital prior to our study team being able to discuss the trial with them (n=11), Of the remaining 102 eligible patients for whom enrollment was offered, 56 declined the trial and 46 were enrolled and randomized to the control (n=23) or intervention (n=23) group (*Figure 1*).

**Figure 1.**
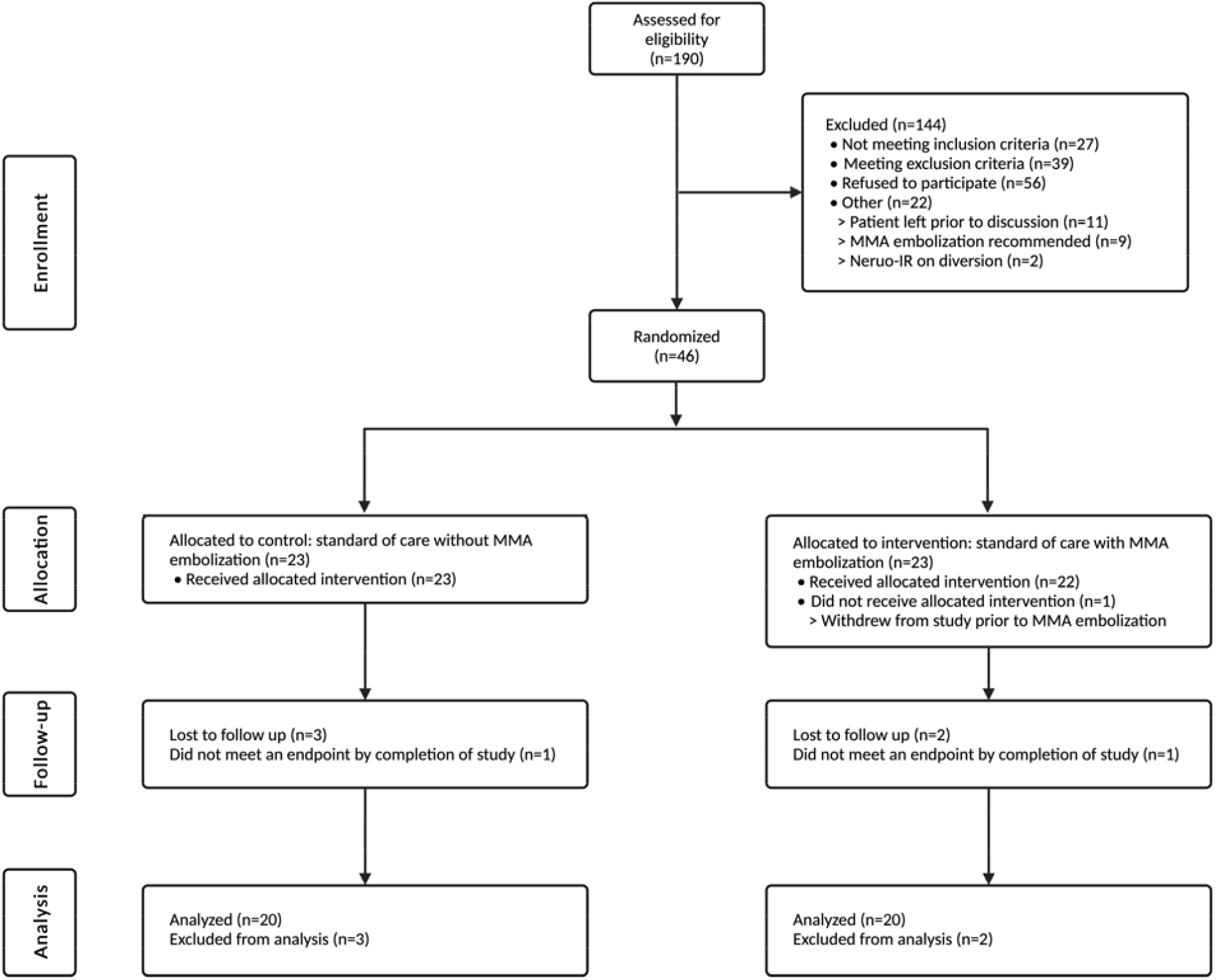
CONSORT Diagram

One patient in the intervention group withdrew from the study after randomization but before receiving either MMA embolization or standard of care surgical drainage. This subject was subsequently removed from the study entirely per the patient’s request and not included in the analysis. Therefore, the intervention group included a total of 22 patients, all of whom received the intended MMA embolization in addition to standard of care. The control group had a total of 23 patients with only 1 receiving MMA embolization due to failure of the standard of care several months into the treatment course.

In July 2023, assessment of the trial progress by investigators revealed that the rate of patient accrual had significantly diminished to the point where it was felt the total enrollment goal could not be reached within a reasonable timeframe. No further patients were enrolled. The last 1-year follow-up of the last subject enrolled was in February 2024.

Of the 23 control subjects only 20 had sufficient follow-up to include in the primary outcome analysis. Of the 23 subjects in the intervention group, one patient withdrew after randomization. Two of the remaining 22 patients in the intervention group had insufficient follow-up to be included in the primary outcome analysis.

### Baseline data

Baseline patient characteristics (*Table 1*) include study subgroup (symptomatic/asymptomatic), sex, age, antiplatelet/anticoagulant use, National Institutes of Health Stroke Scale (NIHSS), modified Rankin Scale (MRS), cSDH laterality/size, and Nakaguchi Classification. Procedure related factors (Table 1) include timing and type of surgical drainage if any, embolization timing and embolic agent.

**Table 1.**
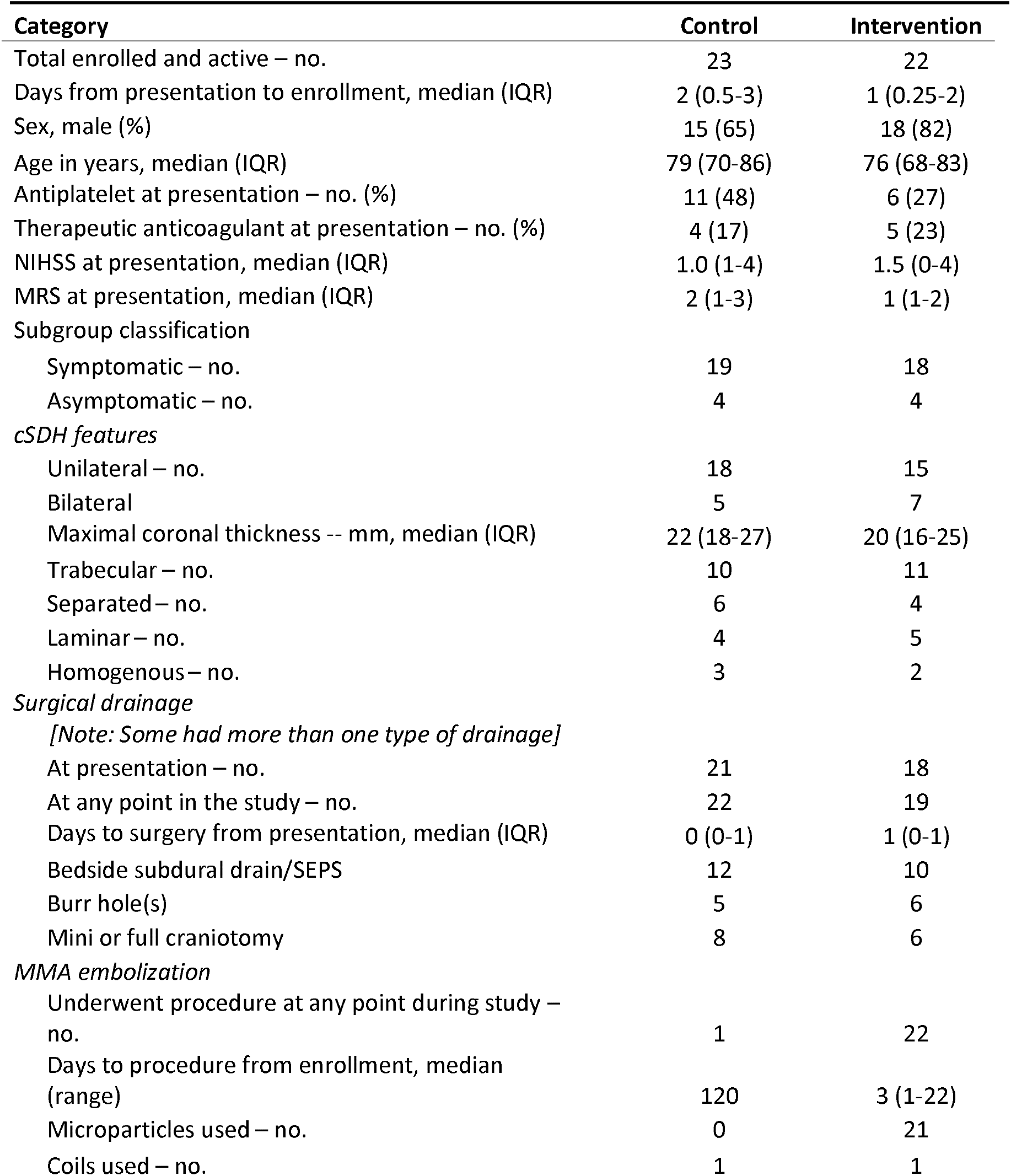
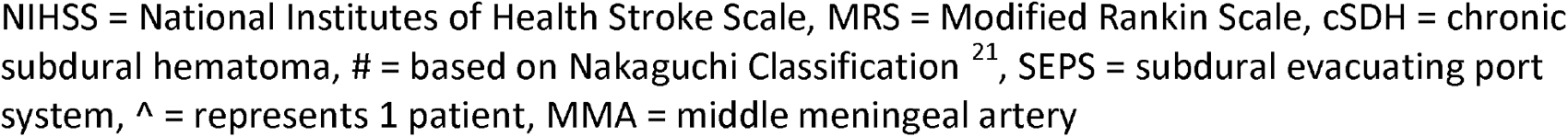
Study Demographics.

| Category | Control | Intervention |
| --- | --- | --- |
| Total enrolled and active – no. | 23 | 22 |
| Days from presentation to enrollment, median (IQR) | 2 (0.5-3) | 1 (0.25-2) |
| Sex, male (%) | 15 (65) | 18 (82) |
| Age in years, median (IQR) | 79 (70-86) | 76 (68-83) |
| Antiplatelet at presentation – no. (%) | 11 (48) | 6 (27) |
| Therapeutic anticoagulant at presentation – no. (%) | 4 (17) | 5 (23) |
| NIHSS at presentation, median (IQR) | 1.0 (1-4) | 1.5 (0-4) |
| MRS at presentation, median (IQR) | 2 (1-3) | 1 (1-2) |
| Subgroup classification |  |  |
| Symptomatic – no. | 19 | 18 |
| Asymptomatic – no. | 4 | 4 |
| *cSDH features* |  |  |
| Unilateral – no. | 18 | 15 |
| Bilateral | 5 | 7 |
| Maximal coronal thickness -- mm, median (IQR) | 22 (18-27) | 20 (16-25) |
| Trabecular – no. | 10 | 11 |
| Separated – no. | 6 | 4 |
| Laminar – no. | 4 | 5 |
| Homogenous – no. | 3 | 2 |
| *Surgical drainage* |  |  |
| [Note: Some had more than one type of drainage] |  |  |
| At presentation – no. | 21 | 18 |
| At any point in the study – no. | 22 | 19 |
| Days to surgery from presentation, median (IQR) | 0 (0-1) | 1 (0-1) |
| Bedside subdural drain/SEPS | 12 | 10 |
| Burr hole(s) | 5 | 6 |
| Mini or full craniotomy | 8 | 6 |
| *MMA embolization* |  |  |
| Underwent procedure at any point during study – no. | 1 | 22 |
| Days to procedure from enrollment, median (range) | 120 | 3 (1-22) |
| Microparticles used – no. | 0 | 21 |
| Coils used – no. | 1 | 1 |

NIHSS = National Institutes of Health Stroke Scale, MRS = Modified Rankin Scale, cSDH = chronic subdural hematoma, # = based on Nakaguchi Classification<sup>21</sup>, SEPS = subdural evacuating port system, ^ = represents 1 patient, MMA = middle meningeal artery

### Outcomes and estimation

The proportion of patients meeting the primary endpoint was similar between the intervention and control groups (*Table 2*). 14 out of 20 patients (70%) in the control group met the primary endpoint while 15 out of 20 patients (75%) in the intervention group met the primary endpoint (odds ratio = 1.3, Fisher’s exact test p-value = 1, 95% confidence interval = 0.3-6.6).

**Table 2.** Outcomes of Trial Patients.

|  | 0-3 months |  | 3-6 months |  | 6-12 months |  | By 1 year |  |
| --- | --- | --- | --- | --- | --- | --- | --- | --- |
|  | Control | Intervention | Control | Intervention | Control | Intervention | Control | Intervention |
| **Primary Endpoints** |  |  |  |  |  |  |  |  |
| Resolution | 8 | 10 | 2 | 3 | 4 | 2 | 14 | 15 |
| Recurrence | 1 | 0 | 1 | 0 | 0 | 0 | 2 | 0 |
| Failed treatment pathway | 2 | 3 | 0 | 0 | 0 | 0 | 2 | 3 |
| Death | 1 | 0 | 0 | 1 | 0 | 0 | 1 | 1 |
| Active (no endpoint) | 9 | 7 | 6 | 3 | 1 | 1 | 1 | 1 |
| Inactive (met endpoint) | - | - | 14 | 15 | 17 | 19 | - | - |
| Lost to follow up (excluded) | 2 | 2 | 0 | 0 | 0 | 0 | 3 | 2 |
| **Total** | **23** | **22** | **23** | **22** | **23** | **22** | **23** | **22** |
| **Included in primary analysis** | - | - | - | - | - | - | **20** | **20** |
| **All cause mortality** | 5 | 0 | 1 | 1 | 1 | 0 | 7 | 1 |
| **Procedural complications** | 4 | 3 | 0 | 0 | 0 | 0 | 4 | 3 |
| Urinary tract infection | 2 | 3 | 0 | 0 | 0 | 0 | 2 | 3 |
| Subdural empyema | 1 | 0 | 0 | 0 | 0 | 0 | 1 | 0 |
| Death | 1 | 0 | 0 | 0 | 0 | 0 | 1 | 0 |
| **NIHSS at follow up** |  |  |  |  |  |  |  |  |
| Median | 0 | 0 | 0 | 0 | 0 | 0 | 0 | 0 |
| Interquartile Range | 1 | 0 | 2 | 0.5 | 3 | 3 | 3 | 3 |
| **MRS at follow up** |  |  |  |  |  |  |  |  |
| Median | 0 | 0 | 0 | 0 | 0 | 0 | 0 | 0 |
| Interquartile Range | 1 | 0 | 3 | 0.5 | 3 | 3 | 3 | 3 |
| **cSDH size at follow up** |  |  |  |  |  |  |  |  |
| Mean | 3.9 | 3.7 | 2.5 | 2 | 1.7 | 1.7 | 1.7 | 1.7 |
| Standard Deviation |  |  |  |  |  |  |  |  |

Control = standard of care (surgical drainage of cSDH or observation), Intervention = standard of care plus middle meningeal artery (MMA) embolization, NIHSS = National Institutes of Health Stroke Scale, mRS = Modified Rankin Scale, cSDH = chronic subdural hematoma

All-cause mortality at 1 year from enrollment was observed in 7 out of 23 patients (30%) in the control group versus 1 out of 22 patients (5%) in the intervention group, which did reach significance (odds ratio = 0.11, Fisher’s exact test p-value = 0.047, 95% confidence interval 0.002-1.0). Only one of the deaths in the control group could be directly attributed to their care for cSDH (see Adverse Events). Of the other deaths in the control group, two were related to stroke or systemic thromboembolic disease, two were unrelated to preexisting disease unrelated to the CNS or thromboembolic disease, and three were of unknown cause.

Details of the clinical and imaging outcomes measures at the 3-, 6-, and 12-month time points are presented in Table 2. No significant differences between control and intervention groups were observed in these measures.

The effects of treatment subgroup (symptomatic vs asymptomatic) were estimated using logistic regression. The rate of treatment failure by MMA embolization and surgical subgroup are shown in Table 3. The results show what could be a clinically significant difference for MMA embolization in the group that was asymptomatic (and therefore not treated with surgical drainage). However, the number of patients in the asymptomatic group was small and the effect of MMA embolization did not reach statistical significance (p=0.052). Nor was the interaction between MMA embolization and surgical drainage significant.

**Table 3.** Rates of failure grouped by MMA embolization and surgical drainage.

| **MMA**<br>**embolization** | **Surgically**<br>**drained** |  |
| --- | --- | --- |
|  | **+** | **-** |
| **+** | 19% (n=16) | 33% (n=3) |
| **-** | 24% (n=17) | 75% (n=4) |

We also performed time-series analysis to assess whether either treatment failure or radiographic cSDH resolution occurred earlier in the group receiving MMA embolization. The log-rank test from Kaplan-Meier analysis showed no significant difference in time-to-failure. Cox regression for resolution of cSDH on CT at 3,6, or 12 months showed no significant difference between the groups.

### Adverse events

In the control group, 4 out of 23 patients (17%) had procedure-related complications: 2 had minor events (2 urinary tract infections) and 2 had major events (1 subdural empyema, 1 death). In the intervention group, 3 out of 22 patients (14%) had procedure-related complications (all minor events, urinary tract infections) (*Table 2*).

## Discussion

We investigated the efficacy of MMA embolization in treatment of chronic subdural hematomas (cSDH) through a single center randomized controlled trial consisting of 46 enrolled patients. We found no difference in our primary outcome within 1 year, defined as resolution of cSDH (size < 5mm) with no return of clinical symptoms, no need for further intervention, and no major treatment-related complications. We also evaluated all-cause mortality as a secondary outcome. We found that patients who were managed with MMA embolization plus standard of care showed significantly lower all-cause mortality at 12 months compared to those receiving standard of care alone. Lastly, we found no quantitative difference in adverse events between these groups.

The need for randomized controlled trials rapidly became evident after the publication of multiple nonrandomized trials which found striking benefit for MMA embolization in cSDH. In a prospective study with a historical control group, Ban et al ^6^enrolled 72 cSDH patients to undergo MMA embolization with and without surgical drainage and reported lower treatment failure (measured as incomplete resolution > 10 mm or need for surgical rescue of symptoms) in those receiving MMA embolization. They found no additional treatment-related complications in the MMA embolization group. A later meta-analysis of nonrandomized trials also showed substantial benefit for MMA embolization with only 2% hematoma recurrence in those treated with the procedure and 28% recurrence in those without ^14^. Adverse events were low, 2.1% in the MMA patients and 4.4% in conventional treatment. A meta-analysis by Chen et al found little difference in hematoma recurrence after embolization when comparing those who had undergone surgical drainage and those who had not ^15^. The rates of recurrence in their study, was higher than the earlier published results at 4-6%. Based on these and similar results, MMA embolization was rapidly incorporated into clinical practice worldwide. Simultaneously, investigators began work on several randomized clinical trials in order to ameliorate the biases likely to be present in the earlier trials ^16^.

In a single center RCT, Lam et al. randomized 36 post-surgical drainage cSDH patient to either MMA embolization or monitoring alone. Similar to our findings, they report no significant difference in symptomatic recurrence requiring surgical drainage, nor in residual hematoma thickness up to 3 months ^17^. Interestingly, they report significantly better functional outcomes at 3 months in the embolization groups measured by modified Rankin Scale. They report no procedural complications.

More recently, Liu et al. completed a multicenter RCT of a much larger number of patients ^18^. They found symptomatic recurrence or cSDH progression in 6.7% of patients treated with MMA embolization vs. 9.9% in the usual care group. Despite the large number of patients, this trial failed to show a statistically significant benefit in cSDH resolution. Of note, their rate of recurrence in the control group was substantially lower than historically reported. They did find a significantly lower incidence of serious adverse events in the treatment group.

Given the magnitude of the treatment effect in the Liu study, the fact that our current study did not observe a statistically significant difference between treatment and control groups, is not difficult to explain. Our study was powered only to reliably show an effect similar to the magnitude of the effect demonstrated in the study by Ban et al. And, owing to slow enrollment, our study did not even meet that accrual goal. Based on the Liu study, the magnitude of the beneficial effect appears to be smaller than that reported in the earlier nonrandomized studies.

Our primary outcome criteria were more stringent than those used in other studies which may further contribute to the lack of an effect. Any patient in our intervention group who had return of clinical symptoms attributable to SDH, or who required re-intervention even in the light of a new injury, was considered to have failed the primary outcome. Choice of primary outcome is particularly critical for multi-faceted disorders such as SDH which presents in older patients, often with multiple co-morbidities, who are prone to repeat injuries. The complex presentation of cSDH also makes it difficult to judge whether persistent or returning symptoms are attributable to a failure of intervention, or rather the broader pathophysiology of the disorder that extends beyond the radiographic hematoma collection.

If one isolates the results to the treatment group, the odds of successful treatment of cSDH with MMA embolization in our trial was lower than that in prior studies. Possible reasons for this discrepancy include differences in patient populations, different outcome criteria, and different interventional techniques. A recent retrospective study of reasons for the failure of MMA embolization identified a 7% clinical failure rate and a 26% radiographic failure rate ^19^. That study identified several factors associated with treatment failure – choice of embolic agent, size of the middle meningeal artery, anticoagulation, and embolization that did not include the MMA trunk. While our general surgical approaches to cSDH drainage are similar to those elsewhere, our MMA technique was largely limited to particle embolization. The choice of embolic materials currently varies widely between institutions worldwide. While many favor particle embolization for the theoretical benefit of deeper small vessel penetration, others favor liquid embolic agents for safety and ease of handling ^20^. While studies to date show similar results for different agents, it is possible that one embolic material may ultimately show better performance than others. The details of particle embolization may also play an important role. Additionally, in our study, the degree of embolization was left to the operating physician; other trials have specified that particular MMA branches be embolized ^18^.

We find an overall all-cause mortality benefit at 12 months for patients who received MMA embolization in addition to the standard of care. This result might suggest a benefit of MMA embolization. However, the relationship is not immediately evident when the details of the deaths are examined. Only one of the deaths occurred as a probable complication of subdural drainage. The overall excess of deaths in the control group may merely reflect the result of chance (despite the randomization in the study). It might reflect some variation in the care received by the two groups given the open nature of the study. However, despite the lack of differences in morphologic behavior of the cSDH between the groups, it might still result from an unknown benefit from MMA embolization. It has been hypothesized that complications linked to surgical drainage or to the withholding of anticoagulant or antiplatelet therapy could serve as a cause of excess morbidity and mortality in patients with cSDH. MMA embolization might therefore have a benefit when it allows care to proceed without surgical drainage or alteration in anticoagulant or antiplatelet regimens.

There were two major complications directly attributable to surgical decompression in our study, one subdural empyema and one death, both occurring in the control group. We found no adverse outcomes associated with the embolization procedure. This result is similar to those from prior nonrandomized studies. Complications from MMA embolization, including vision loss and other forms of stroke, are a well-known possibility, but their incidence has appeared to be very low. Our study, performed with systematic follow-up, confirms these results.

Our study finds that MMA embolization is safe. The patients no adverse events related to the embolization procedure or the use of particle embolic agents. This finding is consistent with previously reported studies, but most of those studies were not prospective. We had a well-defined process for identifying and reporting adverse events, making the safety results robust. Since MMA embolization may be considered an adjunct procedure in patients who undergo surgical drainage, safety is of the utmost importance.

There are important limitations to this study. The most critical limitation of our study was sample size, as discussed above. Further, this study was limited to a single center and is therefore potentially subject to limitations in the variety of patients enrolled and in variety of practice patterns of the treating physicians. Assignments to the treatment subgroups were based on clinical judgements of the attending neurosurgeons, reflecting common practices but subject to local practice preferences. While there is a variety of embolization materials in current use, the embolization material for this study was, with one exception, Embospheres. The number of patients lost to follow-up was small and is unlikely to have substantially influenced the primary outcome result. While the assessment of the primary outcome was performed in a blinded fashion, the patients and assessors were otherwise not blinded to the group assignments.

In conclusion, our additional prospective evidence with careful clinical follow-up indicates that MMA embolization is a safe and well-tolerated procedure in the cSDH patient population. Although further trials are needed to better understand the efficacy of MMA embolization in reducing cSDH recurrence, the evidence is supportive of a potential mortality and functional outcome benefit associated with MMA embolization in addition to the standard of care.

## Data Availability

All data produced in the present study are available upon reasonable request to the authors.

## Acknowledgements

None

## Notes

**Conflict of interest** None

### Competing Interest Statement

The authors have declared no competing interest.

### Clinical Trial

NCT04270955

### Funding Statement

This study did not receive any funding.

### Author Declarations

Ethics committee/IRB of Dartmouth-Hitchcock Medical Center/Dartmouth Health gave ethical approval for this work.

## References

1. Cofano F, Pesce A, Vercelli G, et al. Risk of Recurrence of Chronic Subdural Hematomas After Surgery: A Multicenter Observational Cohort Study. Front Neurol. 2020;11:560269. doi:10.3389/fneur.2020.560269

2. Edlmann E, Giorgi-Coll S, Whitfield PC, Carpenter KLH, Hutchinson PJ. Pathophysiology of chronic subdural haematoma: inflammation, angiogenesis and implications for pharmacotherapy. J Neuroinflammation. 2017;14(1):108. doi:10.1186/s12974-017-0881-y

3. Weigel R, Schmiedek P, Krauss JK. Outcome of contemporary surgery for chronic subdural haematoma: evidence based review. J Neurol Neurosurg Psychiatry. 2003;74(7):937–943. doi:10.1136/jnnp.74.7.937

4. Zhu F, Wang H, Li W, et al. Factors correlated with the postoperative recurrence of chronic subdural hematoma: An umbrella study of systematic reviews and meta-analyses. eClinicalMedicine. 2022;43:101234. doi:10.1016/j.eclinm.2021.101234

5. Kolias AG, Chari A, Santarius T, Hutchinson PJ. Chronic subdural haematoma: modern management and emerging therapies. Nat Rev Neurol. 2014;10(10):570–578. doi:10.1038/nrneurol.2014.163

6. Ban SP, Hwang G, Byoun HS, et al. Middle Meningeal Artery Embolization for Chronic Subdural Hematoma. Radiology. 2018;286(3):992–999. doi:10.1148/radiol.2017170053

7. Shotar E, Mathon B, Rouchaud A, et al. Embolization of the middle meningeal artery for the prevention of chronic subdural hematoma recurrence in high-risk patients: a randomized controlled trial-the EMPROTECT study protocol. J Neurointerv Surg. Published online February 2, 2024. doi:10.1136/jnis-2023-021249

8. Butcher NJ, Monsour A, Mew EJ, et al. Guidelines for Reporting Outcomes in Trial Reports: The CONSORT-Outcomes 2022 Extension. Jama. 2022;328(22):2252–2264. doi:10.1001/jama.2022.21022

9. Hopewell S, Boutron I, Chan AW, et al. An update to SPIRIT and CONSORT reporting guidelines to enhance transparency in randomized trials. Nat Med. 2022;28(9):1740–1743. doi:10.1038/s41591-022-01989-8

10. Moher D, Schulz KF, Altman D. The CONSORT Statement: revised recommendations for improving the quality of reports of parallel-group randomized trials 2001. Explore NY. 2005;1(1):40–45. doi:10.1016/j.explore.2004.11.001

11. Asfora WT, Schwebach L, Louw D. A modified technique to treat subdural hematomas: the subdural evacuating port system. J Med. 2001;54(12):495–498.

12. Tabaddor K, Shulmon K. Definitive treatment of chronic subdural hematoma by twist-drill craniostomy and closed-system drainage. J Neurosurg. 1977;46(2):220–226. doi:10.3171/jns.1977.46.2.0220

13. Hubschmann OR. Twist drill craniostomy in the treatment of chronic and subacute subdural hematomas in severely ill and elderly patients. Neurosurgery. 1980;6(3):233–236.

14. Srivatsan A, Mohanty A, Nascimento FA, et al. Middle Meningeal Artery Embolization for Chronic Subdural Hematoma: Meta-Analysis and Systematic Review. World Neurosurg. 2019;122:613–619. doi:10.1016/j.wneu.2018.11.167

15. Chen H, Colasurdo M, Kan PT. Middle meningeal artery embolization as standalone treatment versus combined with surgical evacuation for chronic subdural hematomas: systematic review and meta-analysis. J Neurosurg. 2024;140(3):819–825. doi:10.3171/2023.7.JNS231262

16. Levitt MR, Hirsch JA, Chen M. Middle meningeal artery embolization for chronic subdural hematoma: an effective treatment with a bright future. J NeuroInterventional Surg. 2024;16(4):329–330. doi:10.1136/jnis-2024-021602

17. Lam A, Selvarajah D, Htike SS, et al. The efficacy of postoperative middle meningeal artery embolization on chronic subdural hematoma - A multicentered randomized controlled trial. Surg Neurol Int. 2023;14:168. doi:10.25259/SNI_208_2023

18. Liu J, Ni W, Zuo Q, et al. Middle Meningeal Artery Embolization for Nonacute Subdural Hematoma. N Engl J Med. 2024;391(20):1901–1912. doi:10.1056/NEJMoa2401201

19. Salem MM, Kuybu O, Nguyen Hoang A, et al. Middle Meningeal Artery Embolization for Chronic Subdural Hematoma: Predictors of Clinical and Radiographic Failure from 636 Embolizations. Radiology. 2023;307(4):e222045. doi:10.1148/radiol.222045

20. Scoville JP, Joyce E, A Tonetti D, et al. Radiographic and clinical outcomes with particle or liquid embolic agents for middle meningeal artery embolization of nonacute subdural hematomas. Interv Neuroradiol J Peritherapeutic Neuroradiol Surg Proced Relat Neurosci. 2023;29(6):683–690. doi:10.1177/15910199221104631

21. Nakaguchi H, Tanishima T, Yoshimasu N. Factors in the natural history of chronic subdural hematomas that influence their postoperative recurrence. J Neurosurg. 2001;95(2):256–262. doi:10.3171/jns.2001.95.2.0256

